# Urinary Antimicrobial Peptides and Cytokines as Biomarkers for Recurrent Urinary Tract Infection in Children and Adolescents

**DOI:** 10.64898/2025.12.29.25343150

**Authors:** Guillermo Yepes, Hancong Chloe Tang, Natalie Holdsworth, Kristin Salamon, Laura Schwartz, Christina Ching, Steve Rust, John David Spencer

## Abstract

**Background:** Recurrent urinary tract infection (rUTI) is a major diagnostic and management challenge. Dysregulated innate immune responses, including antimicrobial peptides and cytokines, may underlie UTI susceptibility. This study investigates whether urinary concentrations of antimicrobial peptides and cytokines differ in children and adolescents with a history of rUTI and whether they can accurately classify rUTI status.

**Methods:** Urine samples were collected from 42 girls and adolescent females with a history of rUTI and 37 matched healthy controls. Concentrations of antimicrobial peptides (alpha-defensins 1-3, beta-defensin 1, cathelicidin, secretory leukocyte protease inhibitor, lipocalin 2, and ribonuclease 7) and cytokines (interleukin-1 beta, interleukin-6, interleukin-8, and tumor necrosis factor alpha) were quantified using enzyme-linked immunosorbent assays. A logistic regression model with variable selection was developed to classify rUTI participants based on urinary biomarkers and clinical factors.

**Results:** Compared to controls, participants with rUTI had lower concentrations of beta-defensin 1, cathelicidin, and ribonuclease 7, and higher concentrations of alpha-defensins 1-3, lipocalin 2, and secretory leukocyte protease inhibitor. Cytokine concentrations, including interleukin-1 beta, interleukin-6, interleukin-8, and tumor necrosis factor alpha, were elevated in the rUTI group. The predictive model demonstrated high accuracy, with an area under the receiver operating curve of 0.97 and a prevalence-adjusted area under the precision-recall curve of 0.94.

**Conclusions:** Girls and adolescent females with rUTI exhibit a distinct urinary immune profile characterized by dysregulated antimicrobial peptides and elevated proinflammatory cytokines. A predictive model integrating these biomarkers with clinical features accurately classified rUTI status, supporting their potential utility as diagnostic tools for pediatric UTI.

## INTRODUCTION

Urinary tract infections (UTI) are among the most common bacterial infections in females across the lifespan – contributing to substantial morbidity and healthcare utilization worldwide [1–5]. Although most individuals experience sporadic infections, up to one-third develop recurrent UTI (rUTI) – defined as ≥2 febrile or symptomatic episodes between 30 days and 24 months after the initial diagnosis [6–9]. rUTI is associated with repeated antibiotic exposure, compromised kidney function, psychological distress, and reduced quality of life [10–13]. Despite its prevalence and impact, clinicians lack tools to reliably identify which patients will progress from isolated to recurrent infections.

In pediatric and adolescent populations, strategies to reduce rUTI include management of bowel and bladder dysfunction, circumcision, screening for congenital kidney and urinary tract anomalies, and evaluation of sexual activity [14, 15]. Although these interventions can lower UTI incidence, they only partially explain recurrence risk. Thus, there is a critical need for biological markers that improve risk stratification beyond these clinical risk factors, enabling clinicians to identify patients most likely to develop rUTI.

Antimicrobial peptides (AMPs) and cytokines are key innate immune mediators that show promise as UTI biomarkers [16]. AMPs such as defensins, cathelicidin, ribonucleases (e.g., RNase 7), secretory leukocyte protease inhibitor (SLPI), and metal binding proteins are produced by the urothelium of the lower urinary tract, kidney tubules, or infiltrating immune cells. They exert direct antimicrobial activity and impaired production has been linked to increased UTI susceptibility [17–23]. Cytokines including interleukin 1-beta (IL-1β), IL-6, IL-8, and tumor necrosis factor alpha (TNFα) amplify local inflammation and recruit leukocytes to eradicate uropathogens. Excessive or prolonged cytokine responses can promote kidney injury, compromise the bladder barrier, and enhance infection risk [24–28]. While increased AMP and cytokine expression has been demonstrated during acute UTI, their expression with rUTI is poorly characterized [16, 29–38].

Here, we evaluate urinary AMPs and cytokines in girls and adolescent females with rUTI as well as UTI-naïve controls. We integrate the concentrations of these immune markers with clinical variables using logistic regression to define signatures that distinguish rUTI status. Our findings identify a distinct urinary immune signature in participants with rUTI and demonstrate strong discriminatory performance in classifying youth with recurrent infections. These insights advance our understanding of host defense in rUTI and highlight biomarker candidates that may inform future diagnostic and prevention strategies in pediatric UTI.

## METHODS

### Study Population and Sample Collection

Human subjects research was conducted in accordance with the principles of the World Medical Association’s Declaration of Helsinki, and the study protocol was approved by Nationwide Children’s Institutional Review Board. Informed written consent was obtained from all patients. For participants under 18 years of age, written parental/guardian consent and patient assent were obtained. Because UTI occurs more frequently in females, enrollment was limited to this population.

Participants were enrolled between 2019 and 2022 through Nationwide Children’s primary care, nephrology, or urology clinics. Non-infected, free-voided urine samples were obtained from two groups: girls and adolescent females without UTI (controls), and participants with rUTI – defined as ≥2 febrile or symptomatic, culture-positive infections occurring between 30 days and 24 months following an initial UTI. Prior UTIs were defined according to American Academy of Pediatrics clinical practice guidelines, with culture confirmation when available, and supplemented by clinician documentation when cultures were not obtainable [39]. Study participants did not receive antibiotic therapy within one month prior to the time of urine collection. Samples were collected when participants were asymptomatic and without evidence of an active culture-positive infection.

Exclusion criteria included pregnancy, institutionalization, known immunodeficiency, use of immunosuppressive medications, malignancy, chronic antibiotic use or antibiotic treatment within 1 month of enrollment, impaired kidney function, and urinary tract anomalies (including hydronephrosis, solitary kidney, renal dysplasia, cystic kidney disease, urinary tract obstruction, or neurogenic bladder, dilating vesicoureteral reflux). Controls had no chronic medical conditions, except that 4 were diagnosed with hypertension and 3 had seasonal allergies. In the rUTI group, 6 patients underwent a voiding cystourethrogram, and all demonstrated grade 2 vesicoureteral reflux or less. Clinical and demographic data were extracted from the electronic health record, and detailed information on all participants is outlined in **Supplemental Tables 1-2**. Urine was collected with Assay Assure (Thermo-Fisher, Waltham, MA, USA), centrifuged, and supernatants were stored at −80°C until analysis. A dipstick urinalysis and culture was performed on all collected urine specimens.

### ELISA

Commercial assays were used to quantify urine concentrations of α-defensins 1-3 (Hycult Biotech, Plymouth Meeting, PA, USA), β-defensin 1 (PeproTech, Cranbury, NJ, USA), cathelicidin (Hycult), lipocalin 2 (Abcam, Cambridge, United Kingdom), RNase 7 (Hycult), and SLPI (Abcam). Assays were performed following manufacturer’s instructions. Urinary cytokines (IL-1β, IL-6, IL-8, and TNFα) were measured using the multiplex V-Plex Viral Panel 1 (Meso Scale Diagnostics, Rockville, Maryland, USA). Urine creatinine was also measured (Oxford Biomedical Research, Rochester Hills, MI, USA).

### Quantification and statistical analysis

Cytokine concentrations were log10-transformed to improve normality. AMP concentrations were reported in absolute concentrations to preserve interpretability. Continuous variables were assessed for normality with the D’Agostino-Pearson Omnibus or Shapiro-Wilk normality test, with normality defined as a *P-*value > 0.05. Group comparisons were performed using a Student’s *t*-test or ANOVA for normally distributed data and the Mann-Whitney *U* or Kruskal-Wallis tests for nonparametric data.

### Prediction model development and evaluation

A supervised machine learning logistic regression model with least absolute shrinkage and selection operator (LASSO) regularization was developed to classify rUTI status based on urinary biomarkers and clinical covariates. Candidate variables included all AMPs, log10-transformed cytokines, urine creatinine, and clinical metrics including age at sample collection, body mass index percentile, and leukocyte esterase (LE). Model training used 10 repetitions of 10-fold stratified cross-validation to identify the optimal predictive feature set and reduce overfitting. Final model coefficients were obtained by fitting the LASSO model on the full data set with the best hyperparameter chosen by repeated cross-validation.

Model performance and uncertainty were estimated using bootstrap resampling (1,000 iterations), with each bootstrap sample undergoing ten repetitions of 10-fold stratified cross-validation using the best hyperparameter value. Performance metrics included area under the receiver operating characteristic curve (AUROC), area under the precision-recall curve (AUPRC), sensitivity, specificity, accuracy and positive predictive value (PPV). PPV and accuracy were calculated using the historical prevalence of rUTI in the target population (25%) rather than the prevalence in the study data set [2]. PPV and sensitivity were plotted as a function of the percentage of individuals intervened to illustrate performance across operational thresholds. All statistical analyses were performed in R (v4.4.2) using the packages *rsample, glmnet, caret, pROC, precrec,* and *boot*.

## RESULTS

### Patient characteristics

We enrolled 79 toilet-trained girls and adolescent females, including 42 with a history of rUTI and 37 controls with no UTI history (**Table 1**). Children with rUTI had a median of 4 prior infections, with uropathogenic *E. coli* (UPEC) being the most common uropathogen. To protect participant confidentiality, individual-level clinical tables are omitted from this preprint and will be provided in the final peer-reviewed publication.

**Table 1.**
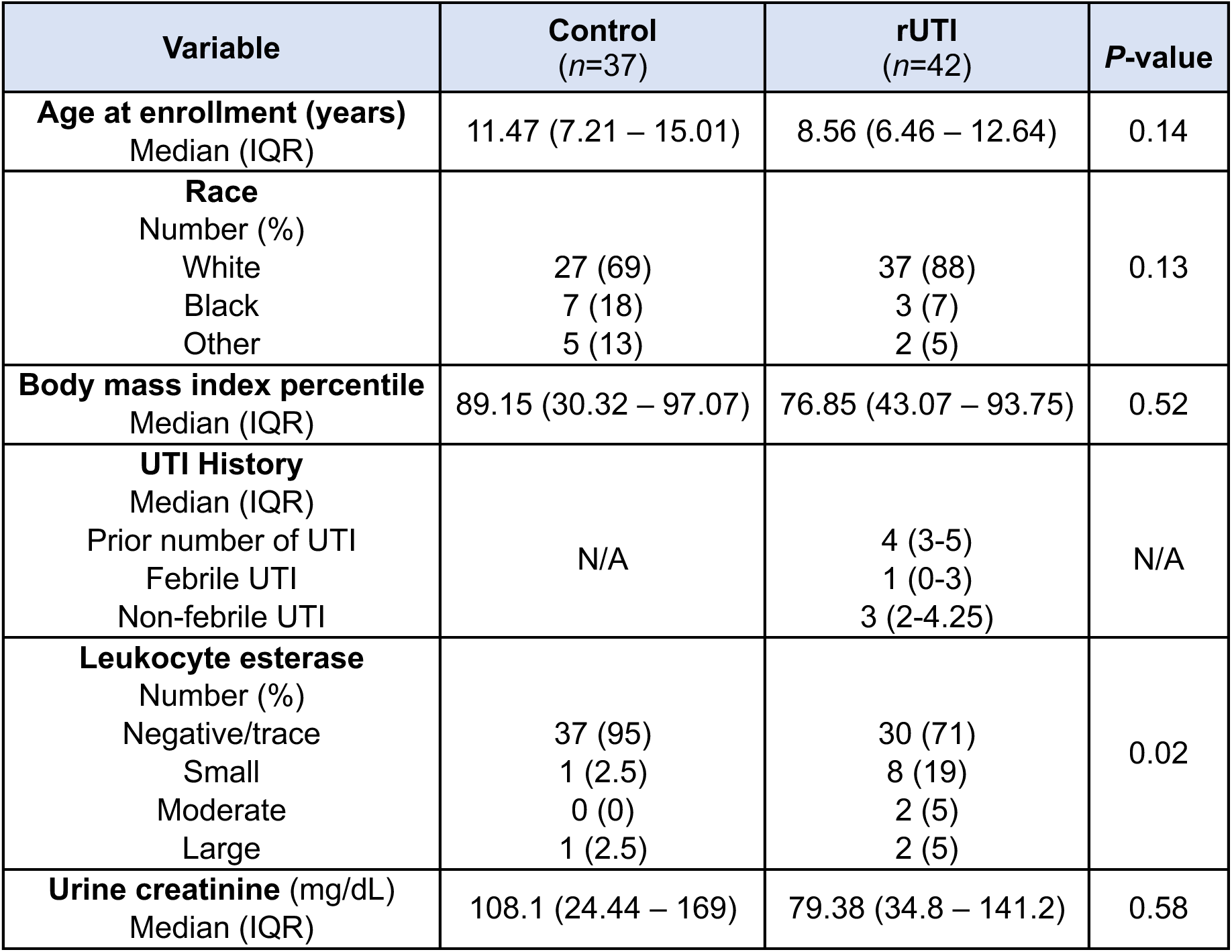
Subject demographics and clinical characteristics. Values are presented as median (interquartile range) or number (percent), as appropriate. *P*-values for age at enrollment, body mass index percentile, and urine creatinine were calculated using the Mann-Whitney *U* test. *P*-values for race and leukocyte esterase were calculated using Fisher’s exact test. N/A indicates variables not applicable to the control group.

| Variable | Control<br>(n=37) | rUTI<br>(n=42) | P-value |
| --- | --- | --- | --- |
| **Age at enrollment (years)**<br>Median (IQR) | 11.47 (7.21 – 15.01) | 8.56 (6.46 – 12.64) | 0.14 |
| **Race**<br>Number (%) |  |  | 0.13 |
| White | 27 (69) | 37 (88) |  |
| Black | 7 (18) | 3 (7) |  |
| Other | 5 (13) | 2 (5) |  |
| **Body mass index percentile**<br>Median (IQR) | 89.15 (30.32 – 97.07) | 76.85 (43.07 – 93.75) | 0.52 |
| **UTI History**<br>Median (IQR)<br>Prior number of UTI<br>Febrile UTI<br>Non-febrile UTI | N/A | 4 (3-5)<br>1 (0-3)<br>3 (2-4.25) | N/A |
| **Leukocyte esterase**<br>Number (%) |  |  | 0.02 |
| Negative/trace | 37 (95) | 30 (71) |  |
| Small | 1 (2.5) | 8 (19) |  |
| Moderate | 0 (0) | 2 (5) |  |
| Large | 1 (2.5) | 2 (5) |  |
| **Urine creatinine (mg/dL)**<br>Median (IQR) | 108.1 (24.44 – 169) | 79.38 (34.8 – 141.2) | 0.58 |

### Urinary antimicrobial peptides are dysregulated with rUTI

We observed two distinct patterns of AMP expression. Peptides primarily produced by the uroepithelium were suppressed, while peptides largely derived from infiltrating leukocytes were elevated [23, 26]. Compared to controls, participants with rUTI had lower urinary concentrations of β-defensin 1 [median (interquartile range)]: [500.6 (436.5 – 552.8) vs. 401.8 (345.3 – 430.8) pg/mL], cathelicidin [8.37 (7.1 – 10.47) vs. 4.9 (3.45 – 7.19) ng/mL], and RNase 7 [445.7 (285.7 – 2559) vs. 236.4 (170.5 – 562.9) ng/mL]. In contrast, α-defensins 1-3 [1005 (263.8 – 3089) vs. 3765 (2559 – 4327) pg/mL], lipocalin 2 [823 (200.4– 1706) vs. 1402 (678.9 – 2227) pg/mL], and SLPI [731.5 (313.6 – 810.4) vs. 1232 (722.5 – 1509) pg/mL] were significantly elevated in the rUTI cohort (**Figure 1**). Normalization to urine creatinine yielded similar trends, although β-defensin 1 no longer reached significance (**Supplemental Figure 1**).

**Figure 1.**
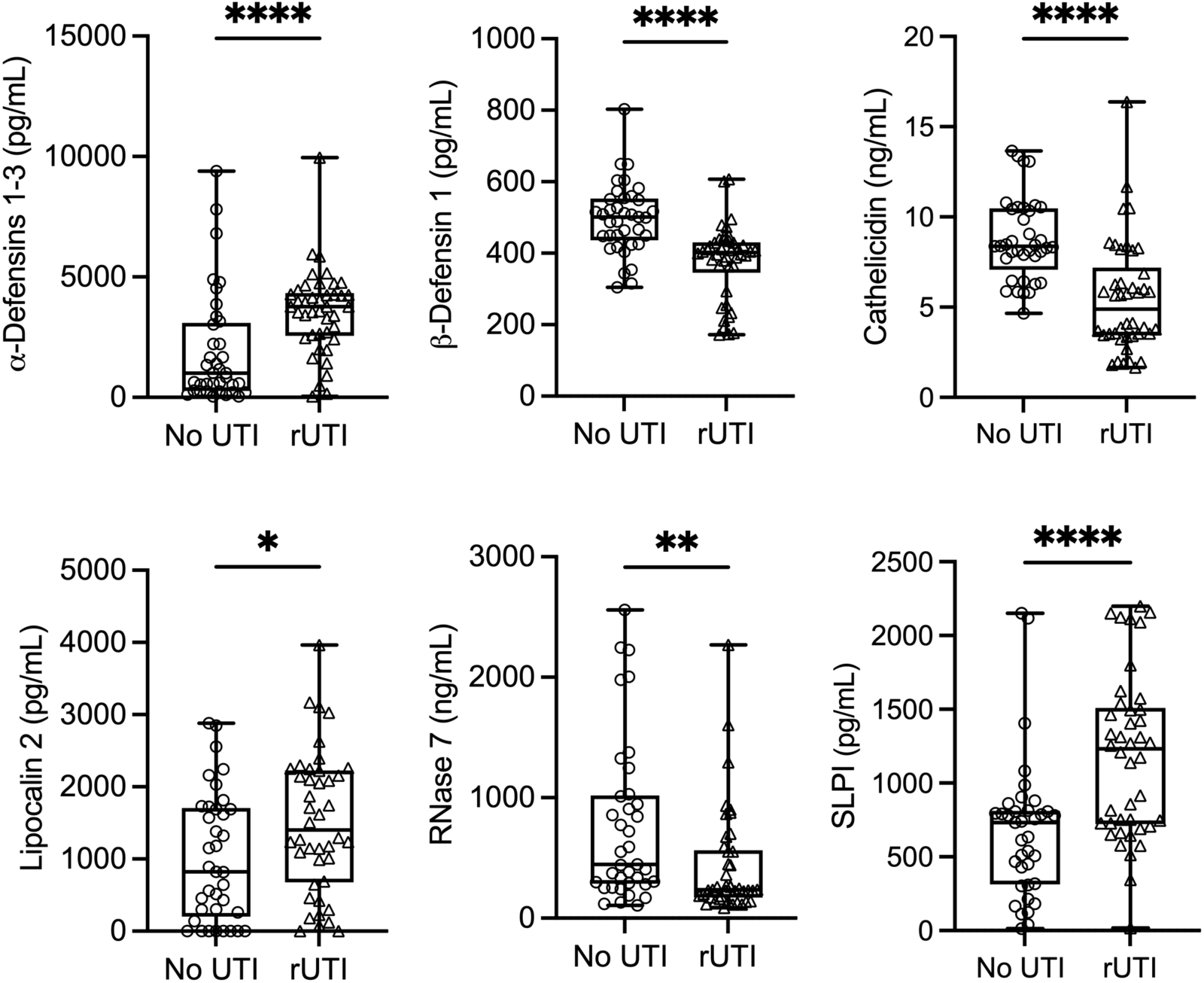
Urine antimicrobial peptides concentrations are dysregulated in girls and adolescent females with rUTI. Urinary α-defensins 1-3, β-defensin 1, cathelicidin, lipocalin 2, RNase 7, and SLPI concentrations in controls (no UTI) and youth with rUTI. Center lines show the median values, box limits indicate the 25^th^ and 75^th^ percentiles, and whiskers show the minimum to maximum range. Each symbol denotes a measurement in a different person. Asterisks denote significant *P*-values for the indicated comparisons (Mann-Whitney *U* test). \**P* <0.05, \*\**P* <0.01, and \*\*\*\**P* <0.0001.

Across the entire cohort, AMP concentrations were stratified by leukocyte esterase (LE) on dipstick urinalysis (negative/trace vs. ≥ small). α-defensins 1-3 and lipocalin 2 were significantly elevated in LE-positive samples, consistent with their leukocyte origin. SLPI showed a similar pattern but did not reach significance. In contrast, β-defensin 1, cathelicidin, and RNase 7 showed no correlation with LE – supporting their uroepithelial origin (**Supplemental Figure 2**). Additionally, the number of prior UTIs showed inverse correlations with epithelial-derived AMPs and positive correlations with leukocyte-associated AMPs, consistent with dysregulation of urinary AMPs in children with more frequent infections (**Supplemental Figure 3**).

### Urinary cytokines are elevated with rUTI

Females with rUTI had higher concentrations of measured cytokines compared to controls. Median IL-1β [0.18 (0.06 – 0.83) vs. 0.51 (0.1 – 2.55) pg/mL], IL-6 [0.18 (0.06 – 0.54) vs. 0.31 (0.16 – 0.78) pg/mL], IL-8 [6.25 (1.15 – 26.41) vs. 28.66 (8.66 – 140.5) pg/mL], and TNFα [0.03 (0.02 – 0.04) vs. 0.04 (0.03 – 0.07) pg/mL] concentrations were elevated in the rUTI cohort (**Figure 2**). Cytokine concentrations normalized to urine creatinine demonstrated similar trends (**Supplemental Figure 4**).

**Figure 2.**
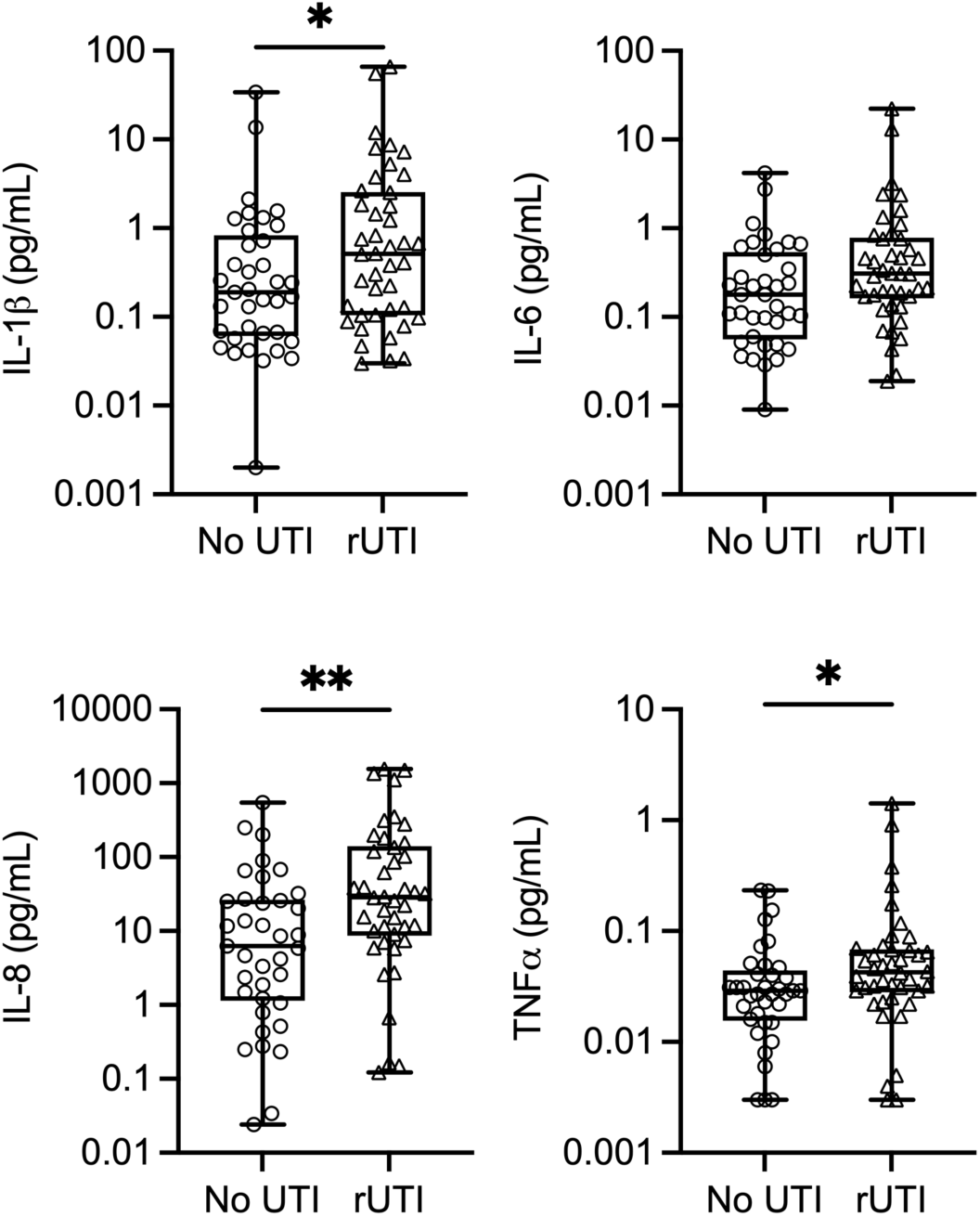
Urine cytokine concentrations are elevated in girls and adolescent females with rUTI. Urinary IL-1β, IL-6, IL-8, and TNFα concentrations in controls (no UTI) and youth with rUTI. Center lines show the median values, box limits indicate the 25^th^ and 75^th^ percentiles, and whiskers show the minimum to maximum range. Each symbol denotes a measurement in a different person. Asterisks denote significant *P*-values for the indicated comparisons (Mann-Whitney *U* test). \**P* <0.05 and \*\**P* <0.01.

Across the entire cohort, cytokine concentrations were further stratified by LE and examined in relation to prior number of infections. IL-1β, IL-6, IL-8, and TNFα each showed positive associations with LE, consistent with their role as inflammation-driven proteins (**Supplemental Figure 5**). In addition, cytokine concentrations showed positive associations with the number of prior UTIs, supporting a link between rUTI and sustained inflammatory signaling (**Supplemental Figure 6**).

### Predictive modeling identifies a urinary signature for rUTI classification

Given the distinct urinary AMP and cytokine patterns observed, we next asked whether these markers could be combined with clinical covariates to accurately classify rUTI status. All measured AMPs, cytokines, urine creatinine, and clinically obtained variables (age, body mass index percentile, and LE) were included as candidate predictors in a LASSO logistic regression. Repeated cross-validation and bootstrap resampling were then used to select an optimal subset of predictors and to estimate model performance. The final optimized model, containing only the selected predictors, distinguished rUTI cases from controls with excellent discriminatory performance – achieving a median AUROC of 0.97 (95% CI: 0.94 - 0.99) and a median prevalence-adjusted AUPRC of 0.94 (95% CI: 0.82 - 0.99) (**Figure 3A/B**).

**Figure 3.**
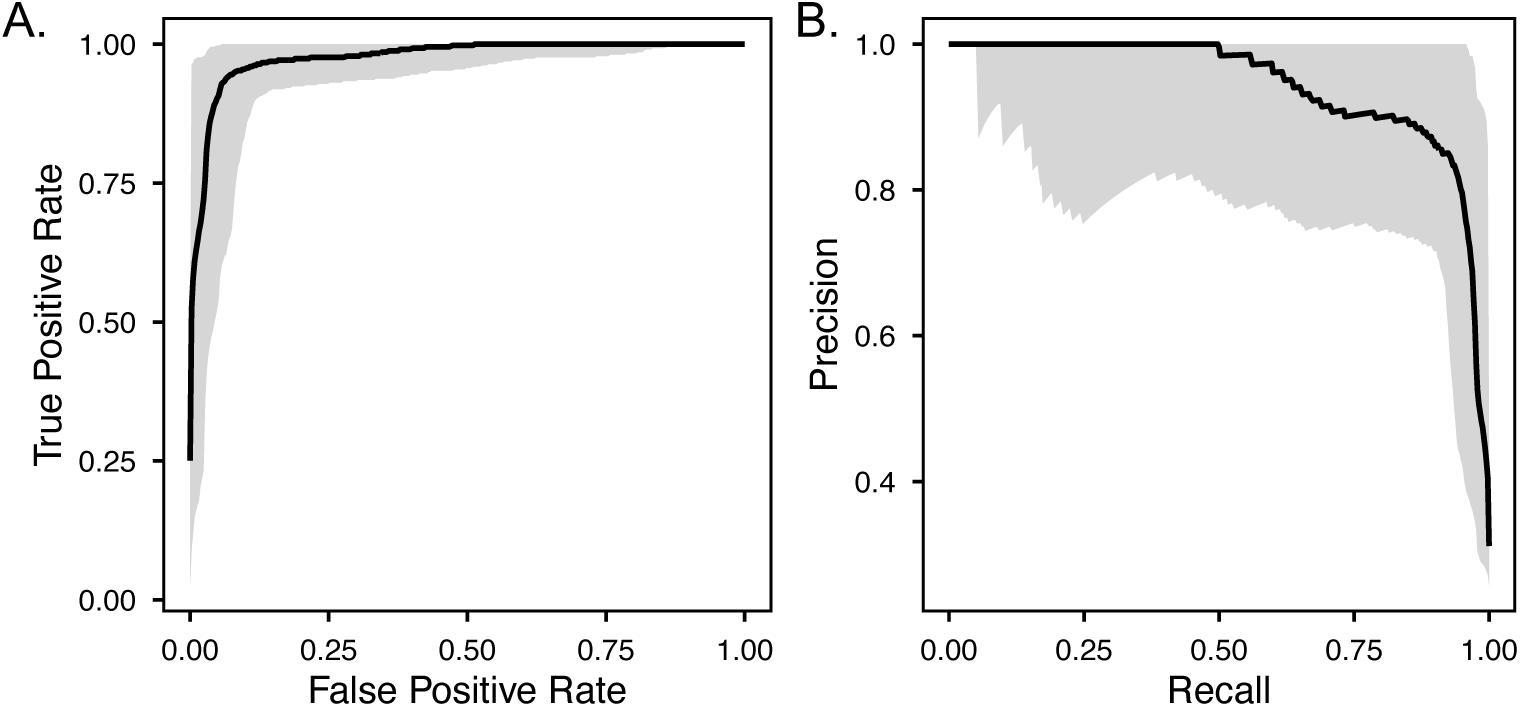
Predictive performance of the LASSO logistic regression rUTI prediction model. (**A**) Receiver operating characteristic (ROC) curve of the logistic regression model integrating urinary antimicrobial peptides, cytokines, and clinical variables. The solid line shows the median ROC curve from 1,000 bootstrapped resamples using repeated cross-validation. The shaded region represents the 95% confidence region derived from these resamples. (**B**) Precision-recall (PR) curve of the same model. The solid line shows the median PR curve based on 1,000 bootstrapped resamples with repeated cross-validation. The shaded region represents the 95% confidence region derived from these resamples.

Threshold analysis demonstrated stable performance across a range of decision boundaries. At the optimal threshold of 0.57 – selected to maximize prevalence-adjusted accuracy (**Figure 4A**) – the model achieved 94.1% accuracy (95% CI: 88.9 - 98.7), 93.6% sensitivity (95% CI: 87.1 – 98.8), and 94.3% specificity (95% CI: 87.8 – 99.5). At this threshold, the median PPV was 84.6% (95% CI: 71.6–98.4), while the negative predictive value was 97.7% (95% CI: 95.7–99.6). To illustrate clinical tradeoffs, PPV and sensitivity were evaluated across increasing proportions of individuals ranked by predicted risk. Intervening on the top 21.9% of individuals yielded a PPV of 100% with a sensitivity of 41.7%, whereas intervening on the top 47.2% achieved a balanced PPV and sensitivity of approximately 85%, highlighting flexibility in selecting operational thresholds based on clinical priorities (**Figure 4B**).

**Figure 4.**
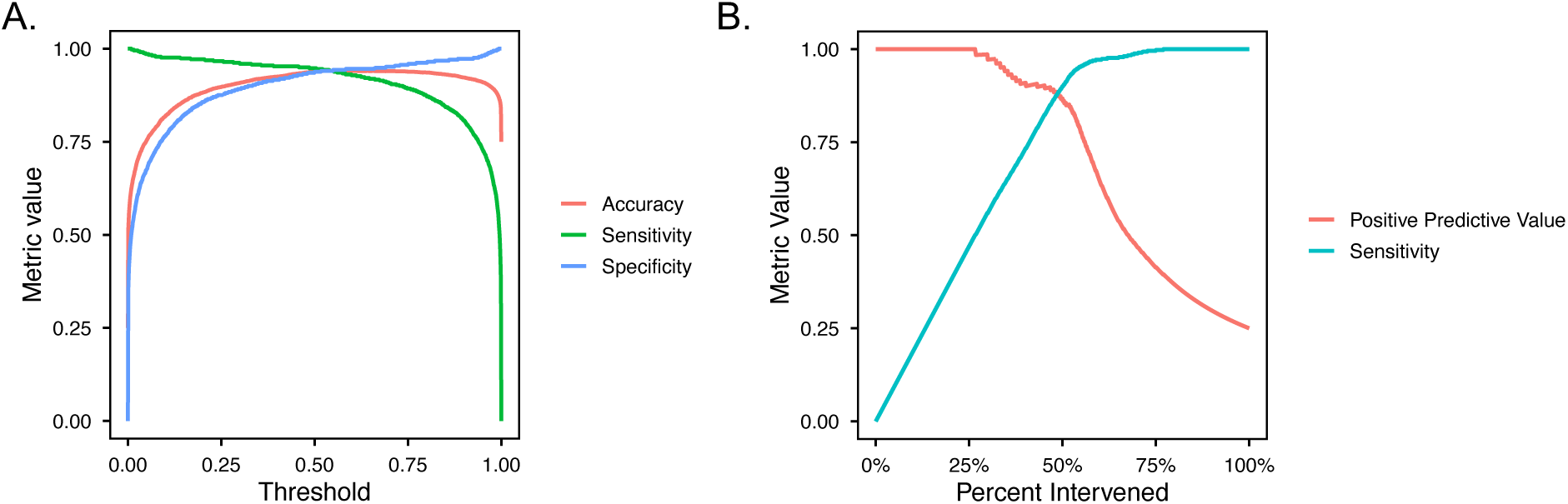
Threshold dependent performance metrics for the LASSO logistic regression rUTI prediction model. (**A**) Sensitivity, specificity, and prevalence-weighted accuracy plotted across rUTI probability thresholds. The vertical line indicates the threshold (0.57) that maximized prevalence-weighted accuracy. (**B**) Prevalence-adjusted positive predictive value (left y-axis) and sensitivity (right y-axis) versus the percentage of individuals the model would flag for clinical action (i.e., predicted to have rUTI) at each risk-score threshold, based on aggregated predicted probabilities from repeated cross-validation. Both curves represent median metric values from 1,000 bootstrapped samples.

Model feature stability was evaluated using bootstrap resampling. A subset of parameters was repeatedly retained across bootstrap iterations, including SLPI, α-defensins 1-3, LE, RNase 7, β-defensin 1, and cathelicidin. In contrast, cytokines, age, body mass index percentile, and urine creatinine were inconsistently selected across bootstrap iterations and were not retained in the final optimized model. The stable predictors were refit to obtain the final model coefficients and odds ratios (**Table 2**), from which the AUROC and AUPRC values were derived. In this model, higher urinary concentrations of SLPI, α-defensins 1–3, and leukocyte esterase were associated with increased odds of rUTI, whereas higher concentrations of RNase 7, β-defensin 1, and cathelicidin were associated with reduced odds of rUTI. These findings suggest that a urinary AMP-dominant signature provides strong discriminatory power for identifying youth with rUTI.

**Table 2.** Final LASSO logistic regression model identifying predictors of rUTI. Regression coefficients (β) were obtained by fitting the LASSO logistic model to the full data set using the optimal penalty parameter selected by repeated cross-validation. Odds ratios (OR) were calculated as the exponential of the regression coefficient (exp β), representing the multiplicative change in the odds of rUTI per unit increase in each predictor. Positive coefficients (OR > 1) indicate association with increased rUTI risk, while negative coefficients (OR < 1) indicate association with reduced risk.

| Feature | LASSO Logistic $\beta$ | Odds Ratio |
| --- | --- | --- |
| SLPI (pg/mL) | 0.57 | 1.77 |
| α-defensins 1-3 (pg/mL) | 0.46 | 1.58 |
| Leukocyte esterase (positive) | 0.36 | 1.43 |
| RNase 7 (ng/mL) | -0.54 | 0.58 |
| β-Defensin 1 (pg/mL) | -0.75 | 0.47 |
| Cathelicidin (ng/mL) | -0.80 | 0.45 |

## DISCUSSION

Key findings from this study demonstrate that girls and adolescent females with a history of rUTI exhibit a urinary immune profile characterized by dysregulated AMPs and elevated cytokines. Epithelial-produced AMPs such as β-defensin 1, cathelicidin, and RNase 7 were suppressed, while leukocyte-expressed AMPs including α-defensins 1-3, lipocalin 2, and SLPI were elevated, alongside higher concentrations of IL-1β, IL-6, IL-8, and TNFα. These abnormalities suggest that impaired urothelial defenses and a heightened inflammatory milieu may predispose individuals to recurrent infections. Further, they highlight the dynamic interplay between epithelial and leukocyte-derived immune responses in shaping UTI susceptibility. Importantly, integration of these biomarkers with clinical features in a supervised machine learning model identified a compact urinary signature that achieved excellent classification accuracy, supporting their potential as rUTI diagnostic tools.

Our finding that cathelicidin and RNase 7 are suppressed with rUTI supports prior preclinical and clinical studies linking these peptides to UTI defense. In mouse models, cathelicidin deletion predisposes to acute UTI, while siRNA-mediated silencing of RNase 7 enhances urothelial UPEC invasion [20, 21]. In humans, urinary cathelicidin levels increase during acute UTI but decrease to below-baseline levels with UTI resolution, suggesting that incomplete recovery may increase recurrence vulnerability [37]. Further, in a separate rUTI pediatric cohort we also found lower RNase 7 concentrations [20]. Although β-defensin 1 was suppressed in our rUTI cohort, its contributions to UTI defense are less defined. *Defb1* knockout mice show increased susceptibility to *Staphylococcus* bacteriuria but not to UPEC infection [40, 41]. Still, lower urinary β-defensin 1 was detected in women undergoing pelvic floor surgery with lower concentrations associated with pre-operative positive urine cultures – suggesting that β-defensin 1 deficiency may promote UTI susceptibility in specific clinical contexts [42].

Leukocyte-expressed peptides were also deregulated in our rUTI cohort. α-defensins 1-3 (human neutrophil peptides), lipocalin 2 (neutrophil gelatinase-associated lipocalin), and SLPI are highly expressed in neutrophils but are also produced by kidney and bladder uroepithelial cells, indicating that their urinary levels reflect combined epithelial and leukocyte contributions [17–19, 22]. Elevated urinary α-defensin 1 (human neutrophil peptide 1) has been reported in adults with chronic pyelonephritis and correlates with leukocyte recruitment [43]. Findings for lipocalin 2 are more variable across studies. Early work noted lower urinary lipocalin 2 in children with rUTI [44]. However, subsequent studies have yielded mixed results – one confirming lower lipocalin 2 concentrations in children with rUTI, while another finding no difference when comparing rUTI with culture-negative urine samples or a first-time UTI. These discrepancies may reflect differences in study design, sample sizes, disparate populations, or assay platform (Western blot vs. ELISA) [38, 44–46]. SLPI, another neutrophil-associated peptide, was elevated in our rUTI cohort. In women, SLPI increases with uropathogen detection, but this response is blunted in individuals with recurrent infection, paralleling our observation that repeated UTI may impair SLPI regulation. Preclinical models demonstrate that SLPI knockout mice experience higher bacterial burden and prolonged bladder inflammation after UTI, consistent with its protective role [18].

In parallel with AMP dysregulation, we observed elevated urinary IL-1β, IL-6, IL-8, and TNFα in individuals with a history of rUTI. During acute infections, UPEC activate Toll-like receptor 4 signaling, triggering cytokine and chemokine release that recruit neutrophils and other immune cells to promote inflammation and bacterial clearance [27]. In both humans and animal models, inflammation correlates with UTI severity and recurrence [47, 48]. Pediatric and adult studies show that urinary cytokines like IL-1β, IL-6, and IL-8 rapidly rise during UTI and may help distinguish cystitis from pyelonephritis [31, 33, 49–51]. Preclinical data further demonstrate that during acute UTI in mice, elevated serum levels of granulocyte colony-stimulating factor, IL-6, and the IL-8 analog keratinocyte-derived chemokine (KC) predict chronic cystitis and subsequent infections [52]. Similarly, excessive or prolonged cytokine activation can be detrimental. Experimental models show that excessive or sustained IL-1β or TNFα contribute to more severe and/or increased susceptibility to recurrent infection [47, 53–55]. Our findings extend these observations by demonstrating that cytokine elevations persist in children with rUTI even when sampled in the absence of acute infection. This pattern suggests a chronic, low-grade inflammatory state that may reduce the urothelium’s ability to neutralize uropathogens. Such sustained inflammation may arise from host genetic factors, quiescent intracellular reservoirs or biofilm-like bacterial communities, or lasting epithelial or epigenetic reprogramming – all processes that impact UPEC susceptibility [25, 26, 47].

Importantly, our logistic regression identifies a set of features that classify rUTI status with high accuracy. These findings reinforce evidence showing that urinary immune signatures can sharpen diagnostic precision and complement clinical risk factors to improve rUTI risk stratification [16, 38, 51]. Notably, although cytokines were consistently elevated in individuals with rUTI, they did not independently improve rUTI classification once AMPs were considered in multivariable modeling, suggesting that inflammatory signals may reflect downstream responses rather than primary determinants of recurrence risk. Earlier recognition of individuals at greatest risk for rUTI has major clinical implications. Timely identification could reduce diagnostic uncertainty and enable proactive management – allowing clinicians to intensify surveillance, optimize antimicrobial prophylaxis or non-antibiotic prevention, target behavioral risk factors, or expedite decisions regarding surgical procedures to correct anatomic anomalies before they lead to more severe or repeated infections. High-risk patients might also benefit from closer follow-up after an acute UTI and early referral to subspecialty care, while lower-risk patients could avoid unnecessary antibiotics or invasive evaluations. Such targeted strategies may improve antibiotic stewardship, improve patient quality of life, reduce healthcare costs, and minimize rUTI sequelae.

While this study shows promise for developing urinary biomarker-based tools to identify rUTI, several limitations should be noted. Our sample size was modest, limiting power for subgroup analyses. Most participants were white, underscoring the need to validate these findings in more racially and ethnically diverse populations. Because enrollment was restricted to girls and adolescent females, generalizability to males, neonates, and adults may be reduced. It is also unclear how urinary tract anomalies such as high-grade reflux or neurogenic bladder influence urinary AMPs and cytokines. Samples were cross-sectional, collected at single time points, and therefore do not capture possible longitudinal changes in AMP or cytokine expression during or between infections. Finally, although regression modeling included internal validation with cross-validation and bootstrapping, external validation in independent cohorts will be essential.

In summary, this study identifies a distinct urinary immune signature in girls and adolescent females with rUTI, characterized by suppressed epithelial-produced AMPs, elevated leukocyte-derived peptides, and persistent cytokines. A predictive model integrating primarily AMP features and LE accurately classified rUTI status, highlighting the promise of urinary biomarkers for UTI risk stratification. Future studies need to validate these biomarkers in larger, prospective cohorts, incorporate clinical-grade assays, and test generalizability across diverse populations. Development of biomarker-based diagnostics could enable earlier identification of individuals at risk for UTI recurrence and guide targeted management strategies.

## Data Availability

All data produced in the present work are contained in the manuscript and will be available upon reasonable request.

## ACKNOWLEDGEMENTS AND FUNDING

We thank Sara Lautzenhiser and Isabelle Coburn for assistance with patient enrollment and Dr. Vidhi Tyagi for cytokine measurement. This work was supported by the National Institutes of Health (NIDDK) under awards R01 DK115737 and T35 DK129192, and by the Nationwide Children’s Innovation Foundation.

## AUTHOR CONTRIBUTIONS

J.D.S. supervised the project and contributed to the design and interpretation of all experiments. C.C., and L.S. managed the IRB and/or contributed to study design, analysis, and interpretation. G.Y., N.H., and K.S. performed the ELISA assays. C.T. and S.R. carried out statistical analysis and regression modeling. G.Y., C.T., and J.D.S. wrote the manuscript with input from all authors.

