## Supplemental Figures for "Urinary Antimicrobial Peptides and Cytokines as Biomarkers for Recurrent Urinary Tract Infection in Children and Adolescents"

**SUPPLEMENTAL FIGURES AND FIGURE LEGENDS**

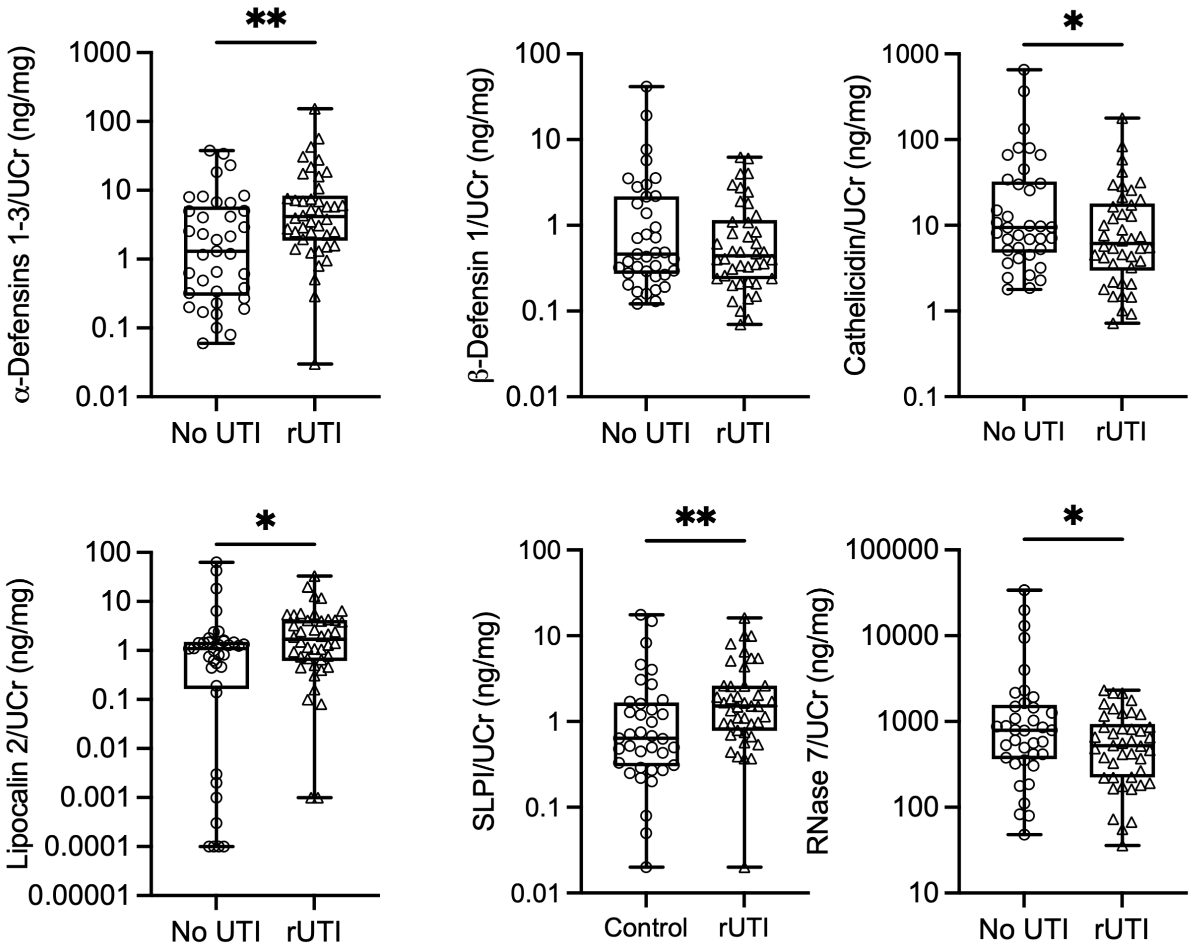


**Supplemental Figure 1. Antimicrobial peptide concentrations normalized to urine creatinine.** Urinary α-defensins 1-3, β-defensin 1, cathelicidin, lipocalin 2, RNase 7, and SLPI concentrations in controls (no UTI) and youth with rUTI normalized to urine creatinine (UCr). Center lines show the median values, box limits indicate the 25$^{th}$ and 75$^{th}$ percentiles, and whiskers show the minimum to maximum range. Each symbol denotes a measurement in an individual participant. Asterisks denote significant *P*-values for the indicated comparisons (Mann-Whitney *U* test). **P* <0.05, ***P* <0.01.

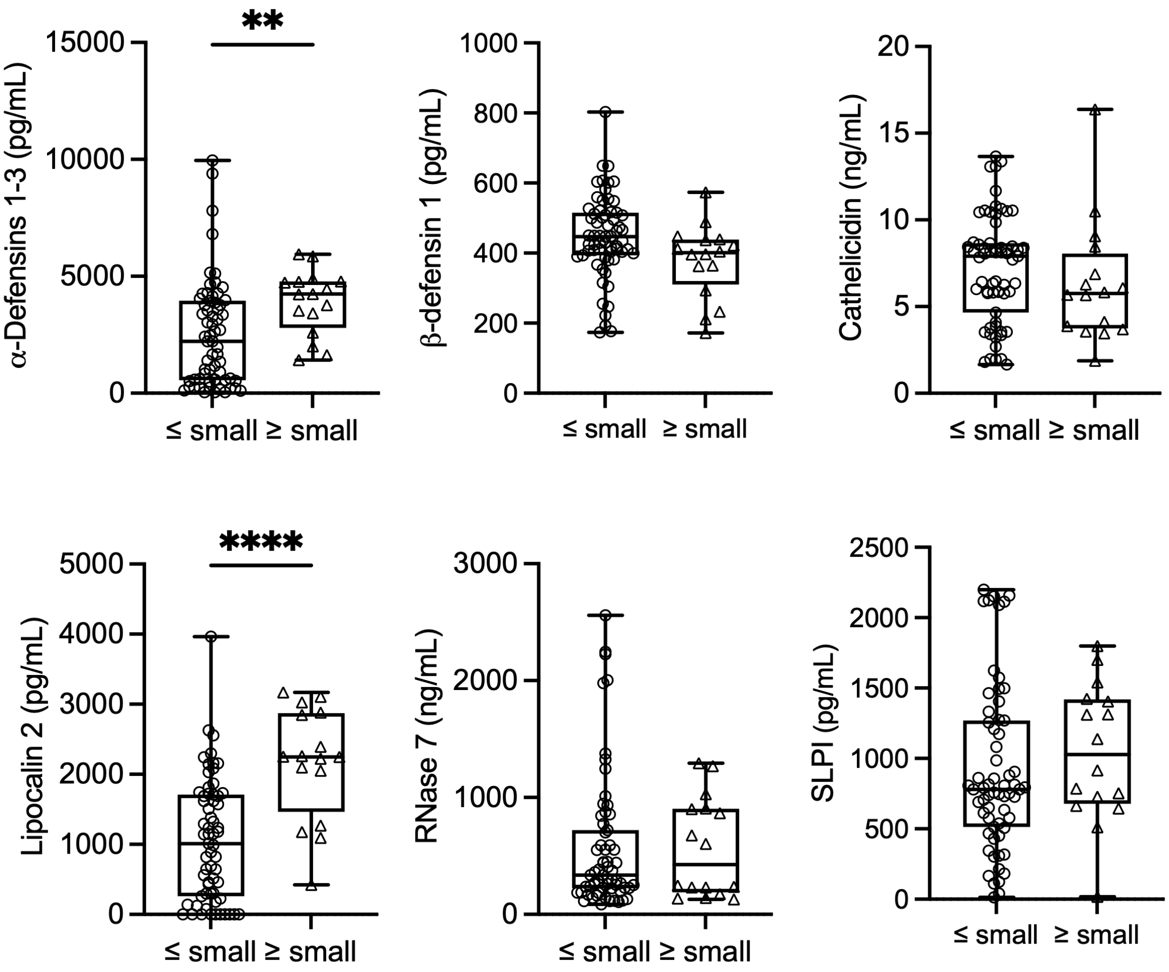


**Supplemental Figure 2. Urine antimicrobial peptides stratified by urine leukocyte esterase.** Urinary α-defensins 1-3, β-defensin 1, cathelcidin, lipocalin 2, RNase 7, and SLPI concentrations in all study participants grouped by urine leukocyte esterase (≤ small (negative/trace) vs. ≥ small) as measured by dipstick urinalysis. Center lines show the median values, box limits indicate the $25^{th}$ and $75^{th}$ percentiles, and whiskers show the minimum to maximum range. Each symbol denotes a measurement in a different person. Asterisks denote significant *P*-values for the indicated comparisons (Mann-Whitney *U* test). ***P* <0.01 and *****P* <0.0001.

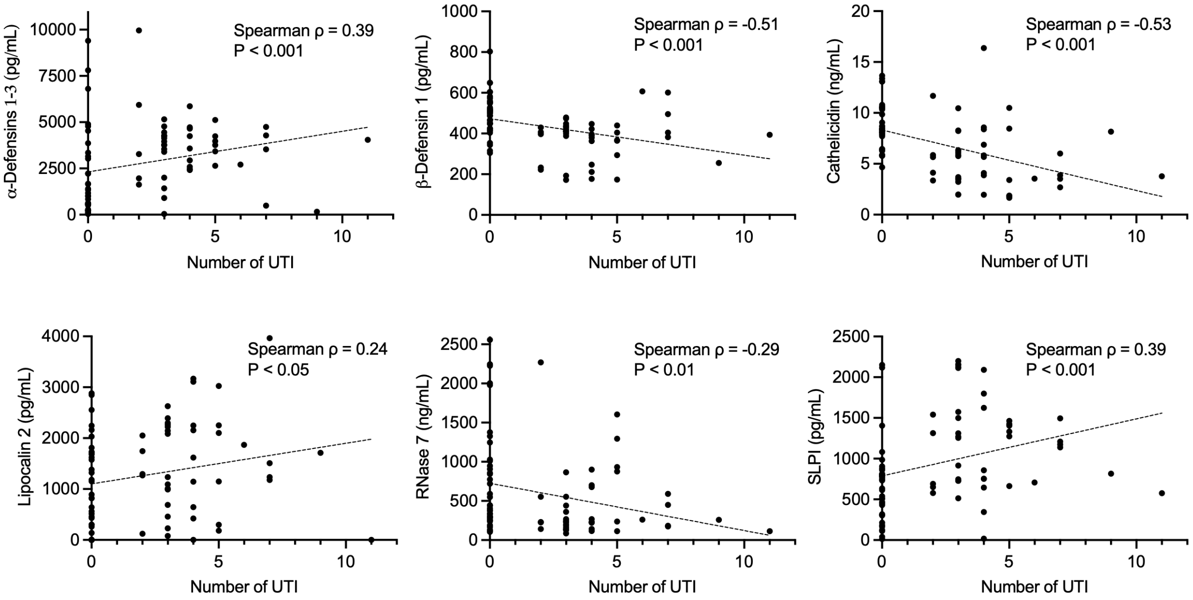


**Supplemental Figure 3. Spearman correlation of urinary antimicrobial peptides with number of prior UTI.** Scatter plots depict the relationship between urinary AMP concentrations and the number of prior UTIs across the entire study cohort. Spearman rank correlation coefficients (ρ) and corresponding two-tailed *P*-values are shown for each AMP. Epithelial-derived AMPs (β-defensin 1, cathelicidin, and RNase 7) demonstrate inverse correlations with UTI number, whereas leukocyte-associated peptides (α-defensins 1–3, lipocalin 2, and SLPI) show positive correlations with increasing UTI burden. Dotted lines indicate linear trend lines for visualization only.

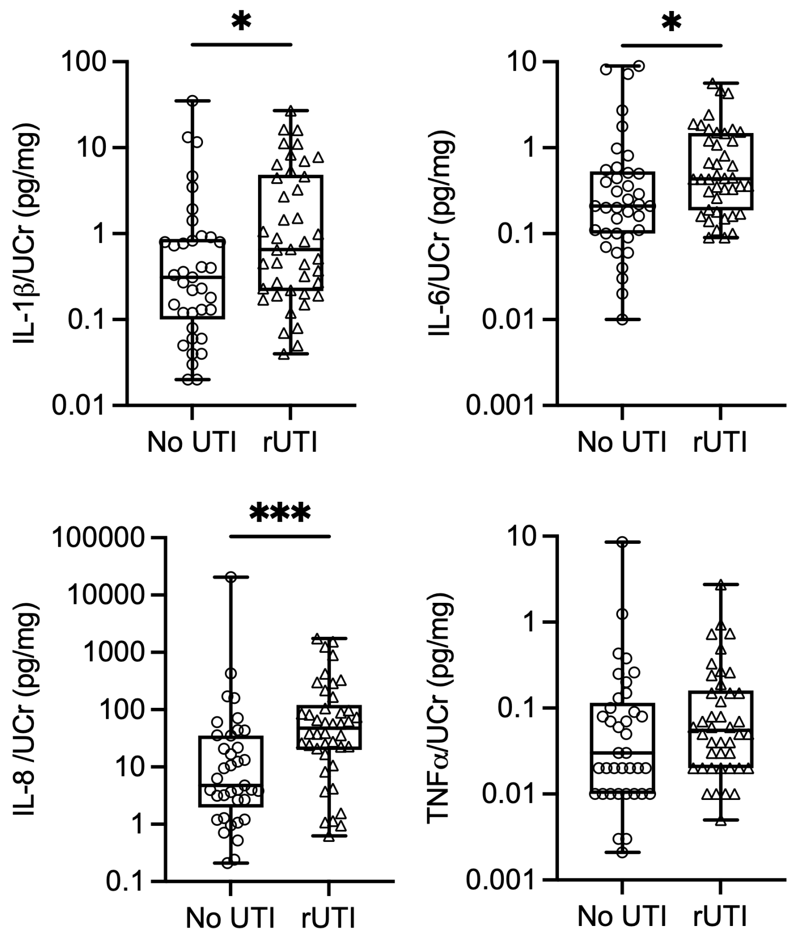


**Supplemental Figure 4. Cytokine concentrations normalized to urine creatinine.** Urinary IL-1β, IL-6, IL-8, and TNFα concentrations in controls (no UTI) and youth with rUTI normalized to urine creatinine (UCr). Center lines show the median values, box limits indicate the $25^{th}$ and $75^{th}$ percentiles, and whiskers show the minimum to maximum range. Each symbol denotes a measurement in a different person. Asterisks denote significant *P*-values for the indicated comparisons (Mann-Whitney *U* test). *$P$ <0.05, ***$P$ <0.001.

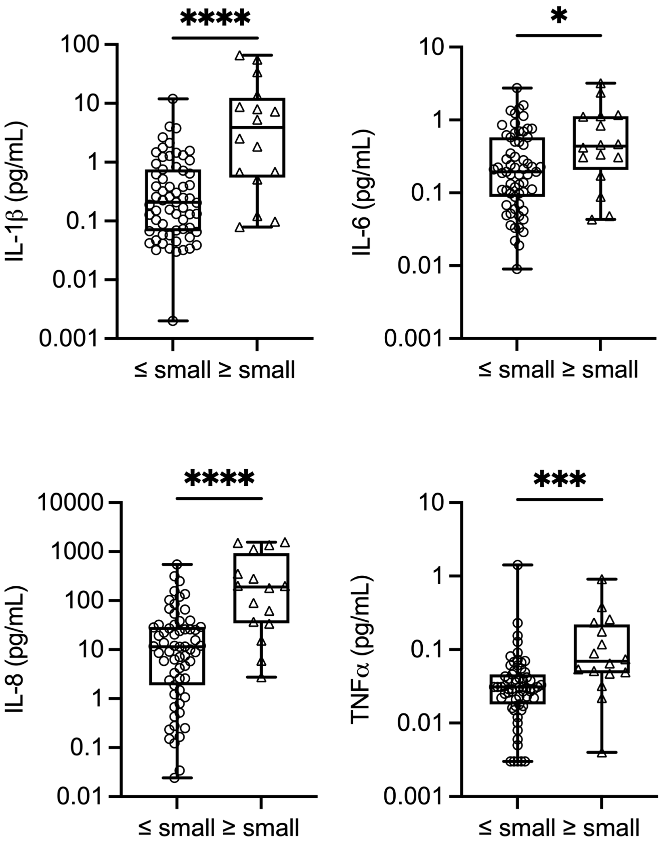


**Suppemental Figure 5. Urine cytokine concentrations stratified by urine leukocyte esterase.** Urinary IL-1β, IL-6, IL-8, and TNFα concentrations in all study participants grouped by urine leukocyte esterase (≤ small (negative/trace) vs. ≥ small) as measured by dipstick urinalysis. Center lines show the median values, box limits indicate the 25$^{th}$ and 75$^{th}$ percentiles, and whiskers show the minimum to maximum range. Each symbol denotes a measurement in a different person. Asterisks denote significant *P*-values for the indicated comparisons (Mann-Whitney *U* test). **P* <0.05, ****P* <0.001, and *****P* <0.0001.

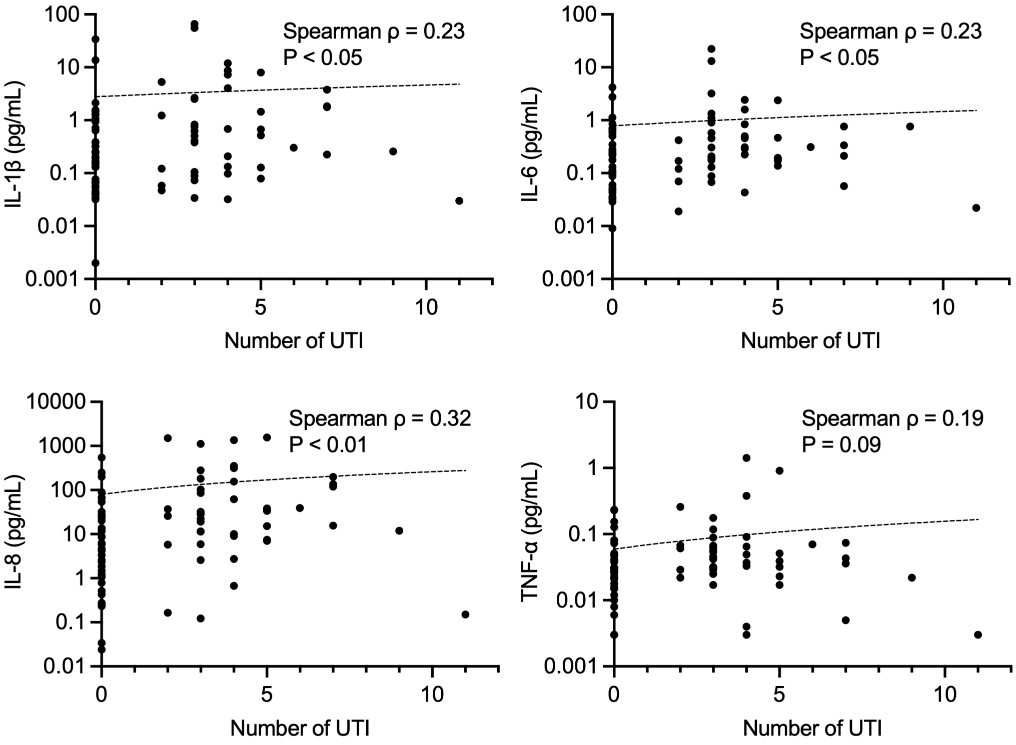


**Supplemental Figure 6. Spearman correlation of urinary cytokines with number of prior UTI.** Scatter plots depict the relationship between urinary cytokine concentrations and the number of prior UTIs across the entire study cohort. Spearman rank correlation coefficients (ρ) and corresponding two-tailed *P*-values are shown for each cytokine. IL-1β, IL-6, and IL-8 demonstrate modest positive correlations with increasing UTI burden, while TNFα shows a weaker, non-significant trend. Dotted lines indicate linear trend lines for visualization only.
